# Childhood Vitiligo: A Retrospective Clinico-Epidemiological Study

**DOI:** 10.1101/2020.09.16.20195636

**Authors:** Ahmed Abdul-Aziz Ahmed, Hayder Saad Ahmed, Mustafa Hameed Mohammed, Mohammed Shanshal

## Abstract

**Background:** Vitiligo is an acquired depigmentary disorder of the skin, mucous membrane and hair follicle resulting from selective destruction of melanocytes.

**Aims of Study:** Identify the clinico-epidemiological characteristics of childhood vitiligo.

**Patients and Methods:** A retrospective study carried out at the dermato-venereology clinic of Salah Al-Din Hospital. A total of 120 vitiligo patients, all younger than 17 years old, were enrolled.

**Results:** Among included patients, (40%) were male and (60%) were female (M:F=2:3). The mean age of onset was (11.5±6.4 years) with (60.8%) of patients were (11-17) years old, (25%) were (6-11) years and (14.2%) were (0-5) years old. Majority of patients were from urban area (87.5%). Generalized types of vitiligo account for (56.7%) besides (22.5%), (17.5%) and (3.3%) represented focal, acrafacial and segmental vitiligo, respectively. Only (8.3%) have nail changes, presented as longitudinal ridging followed by leukonychia. Family history was positive in (37.5%) of vitiligo patients. Treatment used for vitiligo were topical corticosteroids (92.5%), topical calcineurin inhibitors (55%), NB-UVB (84.2%), and systemic steroids (30.8%).

**Conclusions:** Majority of childhood vitiligo develops after puberty and predominantly affects female. Generalized vitiligo is the most frequent type. Atopic dermatitis is the most common associated disease.

## Introduction

Vitiligo is a non-contagious depigmentary disorder that is almost always acquired. It presents as depigmented well defined ivory white macules and patches of different configuration and variable size with unpredictable disease coarse. It may have a generalized, acrofacial, segmental or focal distribution. Selective destruction of melanocytes due to autoimmune mechanisms, intrinsic melanocyte defects or oxidative stress represent possible pathogenic mechanisms.^1^

Worldwide, the prevalence of vitiligo is 1-2% with more than 50% of patients having a childhood onset. Gender distribution among patients shows slight female predominance in childhood vitiligo compared to almost equal prevalence among adults with vitiligo.^2^

Distribution of vitiligo lesions varies among pediatric patients and most frequently presents as generalized disease in 34% followed by acrofacial in 13%, mucosal in 3%, segmental in 29%, and undetermined in 21%.^3^

Childhood vitiligo may cause significant psychological consequences for the patients, causing depression, anxiety, lower self-esteem, and poor quality of life.Anxiety and depression were observed in 26-42% in parents of childhood vitiligo.^4^

Vitiligo is associated with several autoimmune diseases, particularly thyroid disease, pernicious anemia, atopic dermatitis, diabetes mellitus and alopecia areata.^5^ There are several treatment modalities for vitiligo. Topical corticosteroid as currently the first line treatment except for facial and genital vitiligo were topical calcineurin inhibitors are preferred initially. Different types of phototherapy are available with narrow band UVB (NB-UVB) considered safe and highly effective for widespread childhood vitiligo. Systemic immuomodulators and immunosuppressives are used for rapidly progressive disease to achieve rapid control, particularly systemic corticosteroids; however, their use should be limited to a short duration to avoid undesirable side effect.^6^

## Patients and Methods

This work is a retrospective observational study carried out at the outpatient clinic of Salah al- Din Hospital in Iraq. A total number of 120 patients with clinical diagnosis of vitiligo were selected between November 2019 and May 2020. Patients older than 17 years old were excluded.

The age, gender, residence and clinical distribution of vitiligo were recorded for all patients included in the study. Data were analyzed using SPSS version 20. A chi-square and fisher exact test were used to test statistical significance with a P-value of <0.05 was considered as statistically significant.

Regarding ethical consideration, the study was approved by Salah al-Din health directorate, ministry of health, Iraq. Verbal informed consent was taken from all included patients. In addition, all data was kept confidential apart from the purpose of the study.

## Results

Regarding demographic data of patients, females were affected more than males, 60% (n=72) and 40% (n=48), respectively, with a male to female ratio of 2:3 (p-value>0.05). Majority of patients 87.5% (n=105) were from urban region in contrast to 12.5% (n=15) were from rural area. Table 1

**Table 1:** Demographic data of childhood vitiligo.

| **Gender** | **Frequency** | **Percent %** |
| --- | --- | --- |
| **Female** | 72 | 60% |
| **Male** | 48 | 40% |
| **Total** | 120 | 100% |
| **Residence** | **Frequency** | **Percent %** |
| **Urban** | 105 | 87.5% |
| **Rural** | 15 | 12.5% |
| **Total** | 120 | 100% |

The age range was 3-17 years old with the mean age of onset was 11.5±6.4 years. The frequency of patients between 11-17 years old was 60.8% (n=73) with 25% (n=30) of patients were between 6-10 years and only 14.2% (n=17) less than 5 years old. Table 2

**Table 2:** Age of onset of childhood vitiligo in both genders.

| **Age of onset (years)** |  | **Female** | **Male** | **Total** |
| --- | --- | --- | --- | --- |
| **0–5** | No. | 13 | 4 | 17 |
|  | % | 10.8% | 3.3% | 14.2% |
| **6–10** | No. | 18 | 12 | 30 |
|  | % | 15% | 10% | 25% |
| **11–17** | No. | 41 | 32 | 73 |
|  | % | 34.2% | 26.7% | 60.8% |
| **Total** | No. | 72 | 48 | 120 |
|  | % | 60% | 40% | 100% |
| **Pearson Chi-Square= 2.36a** |  | **p-value = 0.306** |  |  |

The study showed that generalized vitiligo 56.7% (n=68) was the most prevalent clinical patten of vitiligo. Focal and acrafacial vitiligo reported in 22.5% (n=27) and 17.5% (n=21), respectively. Segmental vitiligo represented the least common type 3.3% (n=4). In addition, the trunk 46.7% (n=56) was most frequently affected site followed by the head and neck region 33.3% (n=40) while genital and acral sites represented 2.5% (n=3) and 17.5% (n=21), respectively. Table 3

**Table 3:** Patterns and distribution of childhood vitiligo in both genders

|  |  | Gender |  | Total (%) |
| --- | --- | --- | --- | --- |
|  |  | Female | Male |  |
| **Site** | **Head& neck** | 24 | 16 | 40 (33.3%) |
|  | **Trunk** | 31 | 25 | 56 (46.7%) |
|  | **Acral** | 15 | 6 | 21 (17.5%) |
|  | **Genital** | 2 | 1 | 3 (2.5%) |
| **Total** |  | 72 | 48 | 120 |
| Pearson Chi-Square =1.70a p-value= 0.637 |  |  |  |  |
|  |  | Gender |  | Total (%) |
|  |  | Female | Male |  |
| **Types** | **Generalized** | 40 | 28 | 68 (56.7%) |
|  | **Acrofacial** | 12 | 9 | 21 (17.5%) |
|  | **Segmental** | 1 | 3 | 4 (3.3%) |
|  | **Focal** | 19 | 8 | 27 (22.5%) |
| **Total** |  | 72 | 48 | 120 |
| Pearson Chi-Square=3.36a p-value =0.339 |  |  |  |  |

Seventy eight percent (n=84) of patients have no nail findings. However, longitudinal ridging was recorded in 8.3% (n=10) and was most prevalent among females. In contrast, leukonychia was observed in 5% (n=6) which was exclusively reported in males (p<0.05). Table4

**Table 4:** Nail findings in childhood vitiligo.

|  |  | Gender |  | Total (%) |
| --- | --- | --- | --- | --- |
|  |  | Female | Male |  |
| **Nail finding** | **Negative** | 66 | 38 | 104 (86.7%) |
|  | **Longitudinal ridging** | 6 | 4 | 10 (8.3%) |
|  | **Leukonychia** | 0 | 6 | 6 (5%) |
| **Total** |  | 72 | 48 | 120 |
| Pearson Chi-Square = 9.51a p-value =0.008 |  |  |  |  |

Regarding family history of vitiligo, it was negative in 62.5% (n=75) of patients while 37.5% (n=45) reported having a positive family history of vitiligo. In addition, 87.5% (n=105) of patients have no family history of other autoimmune diseases except for psoriasis 5.8% (n=7), thyroid disease 4.2% (n=5) and alopecia areata 2.5% (n=3). Table5

**Table 5:** Family history of autoimmune diseases associated with childhood vitiligo.

|  |  | Gender |  | Total (%) |
| --- | --- | --- | --- | --- |
|  |  | Female (%) | Male |  |
| Family history of vitiligo | Negative | 47 | 28 | 75 (62.5%) |
|  | Positive | 25 | 20 | 45 (37.5%) |
| Total |  | 72 | 48 | 120 |
| Pearson Chi-Square= 0.59a p-value =0.411 |  |  |  |  |
| Family history of autoimmune diseases |  | Gender |  | Total (%) |
|  |  | Female | Male |  |
| Family history | Negative | 64 | 41 | 105 (87.5%) |
|  | Psoriasis | 4 | 3 | 7 (5.8%) |
|  | Thyroid disorders | 3 | 2 | 5 (4.2%) |
|  | Alopecia areata | 1 | 2 | 3 (2.5%) |
| Total |  | 72 | 48 | 120 |
| Pearson Chi-Square = 0.55a p-value =0.813 |  |  |  |  |

Negative past medical history of childhood vitiligo was observed in 75.8% (n=91). Atopic dermatitis was the most frequent associated disorder presented in 7.5% (n=9) followed by autoimmune anemia in 5.8% (n=7) and koebner phenomena in 4.2% (n=5). Each of the following disorders, diabetes mellitus, autoimmune thyroiditis, alopecia areata and halo phenomena, was observed in 1.6% (n=2). Table6

**Table 6:** Past medical history in patients with childhood vitiligo.

|  |  | Gender |  | Total (%) |
| --- | --- | --- | --- | --- |
|  |  | Female | Male |  |
| Past-medical history | Negative | 57 | 34 | 91 (75.8%) |
|  | Diabetes mellitus | 1 | 1 | 2 (1.6%) |
|  | Thyroid disorders | 1 | 1 | 2 (1.6%) |
|  | Autoimmune Anemia | 4 | 3 | 7 (5.8%) |
|  | Alopecia areata | 0 | 2 | 2 (1.6%) |
|  | Atopy | 4 | 5 | 9 (7.5%) |
|  | Koebner phenomena | 3 | 2 | 5 (4.2%) |
|  | Halo phenomena | 2 | 0 | 2 (1.6%) |
| Total |  | 72 | 48 | 120 |
| Pearson Chi-Square = 5.65 a p-value =0.559 |  |  |  |  |

Different modalities were used for the treatment of childhood vitiligo. The study showed 92.5% (n=111) of patients used topical steroids and 55% (n=66) used topical calcineurin inhibitors. UVB was used in 84.2% (n=101). In addition, 30.5% (n=37) of patients used systematic steroids. Females used all types of medical treatments more frequently than males. Table 7

**Table 7:** Treatment modalities for childhood vitiligo in both genders.

|  |  | Gender |  | Total (%) |
| --- | --- | --- | --- | --- |
|  |  | Female | Male |  |
| Topical steroid | Not used | 1 | 8 | 9 (7.5%) |
|  | Used | 71 | 40 | 111 (92.5%) |
| Total |  | 72 | 48 | 120 |
| Fisher's Exact Test= 5.57 a p-value =0.028 |  |  |  |  |
|  |  | Gender |  | Total (%) |
|  |  | Female | Male |  |
| Topical calcineurin inhibitors | Not used | 31 | 23 | 54 (45%) |
|  | Used | 41 | 25 | 66 (55%) |
| Total |  | 72 | 48 | 120 |
| Pearson Chi-Square= 5.22 a p-value =0.880 |  |  |  |  |
|  |  | Gender |  | Total (%) |
|  |  | Female | Male |  |
| NB-UVB | Not used | 16 | 3 | 19 (15.8%) |
|  | Used | 56 | 45 | 101 (84.2%) |
| Total |  | 72 | 48 | 120 |
| Fisher's Exact Test= 3.67 a p-value =0.661 |  |  |  |  |
|  |  | Gender |  | Total (%) |
|  |  | Female | Male |  |
| Systemic steroid | Not used | 48 | 35 | 83 (69.2%) |
|  | Used | 24 | 13 | 37 (30.8%) |
| Total |  | 72 | 48 | 120 |
| Pearson Chi-Square= 0.103a p-value =0.748 |  |  |  |  |

## Discussion

Vitiligo is considered the most common acquired cutaneous depigmentary disease with unpredictable course that is usually associated with significant psychological consequences.^1^ Different studies have demonstrated the increased risk of several autoimmune diseases in patients with vitiligo, particularly diabetes mellitus, thyroid disorders, alopecia areata, psoriasis, and pernicious anemia.^5^

Despite vitiligo was reported to affects both genders in equal proportions in adults and sometimes in children ^7^, childhood vitiligo tends to affect females more frequently than males.^6^ A study by El-Husseiny R et al. 2020 ^8^ involving 483 child with vitiligo observed that females were affected in 59.1% (n=130) of patients. In addition, similar results were observed by Shajil et al. 2006 ^9^ and Nunes & Esser 2011 ^10^. These findings agree with results observed in this study were females represented (60%) of patients. Females are more emotionally fragile to cosmetically disfiguring and socially stigmatizing nature of vitiligo than males. Nevertheless, the role genetic factors and environmental triggers, in addition to the influence of female hormones, should also be considered as possible causes of female dominance.

Several studies have subdivided childhood vitiligo into different age groups. The most common age group reported in this study was 11-17 years old. Kanwar et al. ^11^ 2005 and Brazzelli V et al^12^. 2005 made similar observations. This indicate that more than half of childhood vitiligo develop after puberty which is possibly related to the hormonal changes during this period, particularly in females. However, a study by Gupta M. et al^13^. in 2018 involving 50 patients with childhood vitiligo reported (5.6 years)as the mean age of onset.

Vitiligo presents clinically in different patterns. A study by Ali A. et al.^14^ 2011, enrolling 38 child with vitiligo, recognized generalized vitiligo 44.7% (n=17) as the most common type followed by acrofacial 26.3% (n=10), focal 26.3% (n=10), and segmental vitiligo 2.6% (n=1), an observation consistent with the results of Liu JB. et al^15^. 2005 and Zhang Z. et al^16^. 2009. In this study, results showed that generalized vitiligo 56.7% (n=68) and segmental vitiligo 3.3% (n=4) agree with the above mentioned studies; however, focal vitiligo 22.5% (n=27) was more common than acrofacial vitiligo 17.5% (n=21).

Different studies are contradicted regarding the most common sites affected by vitiligo. The results of this study revealed that the trunk is most common affected site 46.7% (n=56), followed by the head and neck region in 33.3% (n=40) of patients. The acral sites were affected in 17.5% (n=21) with only 2.5% (n=3) of patients have genital lesions. These results contradicted with a study conducted by Ranaivo Irina, et al^17^. 2019 involving 64 child with vitiligo (24.7% in head and 19.1% in trunk). In addition, Sheth et al. 2015^18^ reported that the lower limbs was most frequently involved (62%) followed by the face (46%), the upper limbs (30%), the scalp (25%), and the mucosa (18%).Similar observations were also made by Gupta M. et al^13^. 2018.

We reported nail lesions to occur in 13.3% (n=16) of patients presenting as longitudinal ridging 8.3% (n=10) and as leukonychia 5% (n=6). Topal IO et al^19^. 2016 conducted a case control study enrolling 100 patient with vitiligo, has also observed longitudinal ridging as the most common nail abnormality 42% (n=42) associated with vitiligo (p value=0.001). In addition, he reported that more than two third 78% (n=78) of patients has atleast one nail abnormality. However, his study involved vitiligo in all age group.

The incidence of positive family history in vitiligo patients is 11-46% as reported by different studies^20^. In the current study, positive family history of vitiligo was observed in 37.8% (n=45) of childhood vitiligo. A similar observation was made by Lahlou A. et al. 2017^21^ who reported 39% (n=12) of patients have a family history of vitiligo. A positive family history highlights the role of genetic factors in developing vitiligo, particularly in this country, due to the high incidence of consanguineous marriage.

The prevalence of associated autoimmune diseases in childhood vitiligo are variable among different studies. Autoimmune thyroid disorders are the most commonly reported in association with vitiligo (12.9%)^5^. In contrast, the present study revealed that atopic dermatitis is the most common associated disease 7.5% (n=9) followed by anemia 5.8% (n=7). In addition, alopecia areata was observed in 2.5% (n=2) of this study which is more than double what was observed by Al-Mutairi N et al.22 in 2005 were alopecia areata reported in 1.1% (n=1), and by Handa S et al. 23 in 2003 were 1.3% (n=2) of patients have alopecia areata. Koebner phenomena, as a marker of disease activity, was reported by Agarwal S et al^3^. in 2013 to occur in 24.3% (n=66) of patients with childhood vitiligo compared with 4.2% (n=5) in the present study.

Topical corticosteroids are are considered the first line treatment for limited vitiligo, <20% of body surface area^6^, explaining its use by 92% (n=111) of patients in this study. Several studies have shown that topical corticosteroids is moderately effective to induce repigmentation of vitiligo+24. The high response rate, ease of application, high rate of compliance, and low cost are the advantages of topical corticosteroid therapy for vitiligo. On the other hand, topical calcineurin inhibitors are particularly useful and effective treatment for facial and genital lesions to avoid the side effects of corticosteroids in these regions. Studies by Silverberg et al^25^ 2004 and by Kanwar et al^26^ 2004 demonstrated the efficacy of topical tacrolimus use for vitiligo. It was used by 55% (n=66) of children in the present study.

NB-UVB therapy offers the advantage of being safe in children, no need for photosensitizing agent, and not associated with ocular complications. NB-UVB was used in 84% (n=101) of patients in this study. Kumar Y H et al^27^. 2015 have demonstrated the efficacy of NB-UVB in childhood vitiligo with 76.8% (n=22) of patients achieved complete repigmentation and 14.2% (n=4) achieved partial repigmentation among a total of 28 patients.

Systemic therapy by short course of steroid may be required for rapidly progressive vitiligo either as oral or parenteralroutes^6^. In this study, only 30% (n=37) of patients used systematic steroids. A study conducted by Majid I et al^28^. 2009 demonstrated that mini-pulse oral methylprednisolone is effective in halting the progression of vitiligo in 90% of patients with rapidly progressive disease.

There are few limitations to this study. The duration of this study was relatively shorts to collect large sample size which considered relatively small. In addition, being a single center study is also considered a limitation. However, due to the pandemic of COVID-19, the lockdown of the city and shifting most medical facilities against corona virus, patients faced difficulties access medical care for their vitiligo.

## Conclusion

Childhood vitiligo affects female more than males. The generalized pattern is the most frequent type and mostly affects the trunk. Atopic dermatitis was the most common associated disease while longitudinal ridging is the predominant nail findings in childhood vitiligo.

## Data Availability

The manuscript is available through google drive sharing link.

https://docs.google.com/file/d/1hMLIHKyn5Awatn9eBlJL1RrKgt0ndRHV/edit?usp=docslist_api&filetype=msword

## Compliance with Ethical Standards

### Funding

No funding was received from any source.

### Ethical Approval

- All procedures performed were in accordance with the ethical standards of the institutional and/or national research committee and with the 1964 Helsinki declaration and its later amendments or comparable ethical standards.
- This article does not contain any studies with animals performed by any of the authors.

### Informed Consent

Informed consent was obtained from all individual participants included in the study.

